# Beyond the Haematoma: Socioeconomic Deprivation and Outcomes in Chronic Subdural Hematoma in a Single-Centre Cohort Study

**DOI:** 10.64898/2026.10.07.26364832

**Authors:** Emma Cook, Monica Fisk, Chloe Raggett, Kathryn Bush, Justin Boylan-Toomey, Michael G Hart

## Abstract

**Background:** Chronic subdural hematoma (CSDH) is common and increasing in incidence. Short-term post-surgical outcomes are good, but longer term, patients experience increased morbidity and early mortality than controls and the general population. Deprivation and ethnicity affect many health conditions, but to our knowledge, this is the first UK study investigating their association with CSDH outcomes.

**Methods:** This single-centre retrospective cohort study used Secondary Uses Service Admitted Patient Care Spells (SUS+ APCS) data. Adult subdural hematoma (SDH) patients admitted to St George’s Hospital, London treated with burr-hole surgery between April 2022 and April 2025 were included. Primary outcomes were initial length of stay (LoS) and days spent hospitalised in the subsequent year. Deprivation was measured using Index of Multiple Deprivation (IMD). Linear regression models estimated initial LoS and subsequent days spent hospitalised by category, adjusted for factors including age, ethnicity and comorbidities.

**Results:** In this single-centre cohort of 449 patients, IMD was not associated with initial LoS, but was significantly associated with subsequent time spent hospitalised. In a log-transformed linear regression model, the most deprived spent 4.5-times longer hospitalised during the year after CSDH than the least deprived (fold change = 4.45, 95% confidence interval (95%CI) 2.20 to 9.01).

**Conclusions:** Higher neighbourhood-level deprivation was associated with time spent hospitalised following CSDH. This was robust to adjustment for potential mediating factors. This is the first UK study demonstrating health inequalities in CSDH outcomes, highlighting a need for future research to gain further insights, and to improve equity of outcomes after CSDH.

**What is already known on this topic?:** CSDH is a common sentinel health event, and risk factors include older age, alcohol misuse and taking anticoagulant or antiplatelet medications. Risk factors for poor outcomes include worse pre-morbid functional status and features of more severe CSDH. Neighbourhood-level socioeconomic deprivation is known to be associated with poor health outcomes in many conditions, including fractured neck of femur and stroke.

**What this study adds?:** This is the first UK study to identify that neighbourhood-level deprivation is associated with poorer outcomes after CSDH. We show that greater deprivation is associated with more days hospitalised during the 12 months after CSDH, indicating that the longer-term negative impacts experienced after CSDH may be exacerbated by neighbourhood-level socioeconomic deprivation.

**How this study might affect research, practice or policy:** Future research should replicate these analyses in other regions, and investigate the underlying causal factors underling the inequalities in readmission for CSDH patients. Policy should focus on early intervention to mitigate risk factors for readmission, paying particular attention to those in the most deprived populations.

## INTRODUCTION

Chronic subdural hematoma (CSDH) is among the most common neurosurgical conditions, characterised by the persistent accumulation of blood and inflammatory products within the dura [1]. Risk factors include older age, male sex, having significant comorbidities, alcohol misuse disorder, and taking anticoagulant or antiplatelet medications [2,3]. Globally, CSDH has an incidence of 1.7-20.6 per 100,000 persons per year, and in the UK and Europe its incidence is rising, driven by ageing population structures [4–7]. Surgery remains the mainstay of treatment [8]. CSDH currently costs the UK national health service (NHS) in excess of £42 million per year, projected to rise two-fold by 2040 [9].

While CSDH patients typically have good short-term outcomes, it is recognised as a sentinel health event, with parallels to fractured neck of femur in the elderly [10–13]. CSDH leads to worse long-term mortality and morbidity, with poorer functional, cognitive and mental health outcomes than both controls and the general population [14–18]. Several factors have been identified to influence outcomes after CSDH, including hematoma characteristics, surgical technique, pre-existing comorbidities, and pre-morbid functional status [13,17,19,20].

Socioeconomic deprivation predicts many adverse health outcomes, and impacts overall life expectancy and healthy life expectancy across the globe [21]. In England, neighbourhood-level socioeconomic deprivation, measured using the Index of Multiple Deprivation (IMD) [22], has been shown to negatively affect outcomes after numerous surgical and non-surgical conditions, including cardiovascular disease [23], stroke [24], fractured neck of femur [25],epilepsy [26] and traumatic brain injury [27]. It’s association with outcomes after CSDH, however, remain poorly understood.

To our knowledge, this is the first UK study to investigate the association between neighbourhood-level deprivation and outcomes after CSDH. In this retrospective, single-centre cohort study of surgically managed CSDH patients, we aimed to investigate the impact of IMD on length of stay during the acute admission for CSDH, and the number of days subsequently spent admitted to hospital during the following 12 months.

## METHODS

### Study Design and Patient Population

This study was designed as a single centre retrospective cohort study. All adult subdural hematoma (SDH) patients admitted to St George’s Hospital, London, and treated with burr hole surgery between April 2022 and April 2025 were included. Deidentified data were aggregated from the Secondary Uses Service Admitted Patient Care Spells (SUS+ APCS) database. We used a combination of International Classification of Diseases, Tenth Revision (ICD-10) codes, shown in Figure 1, designed to identify CSDH with maximum sensitivity and specificity. CSDH has no specific ICD-10 code, but the majority of patients are treated with burr hole surgery, whereas acute SDH patients tend to be managed using a craniotomy procedure [28–30]. Patients were excluded if they were under 18 years old, had missing or unclear CSDH aetiology, or were missing an ID number. Any subsequent SDH episodes identified within the timeframe were also excluded, as we were unable to specify whether these were recurrences or new events.

**Figure 1:**
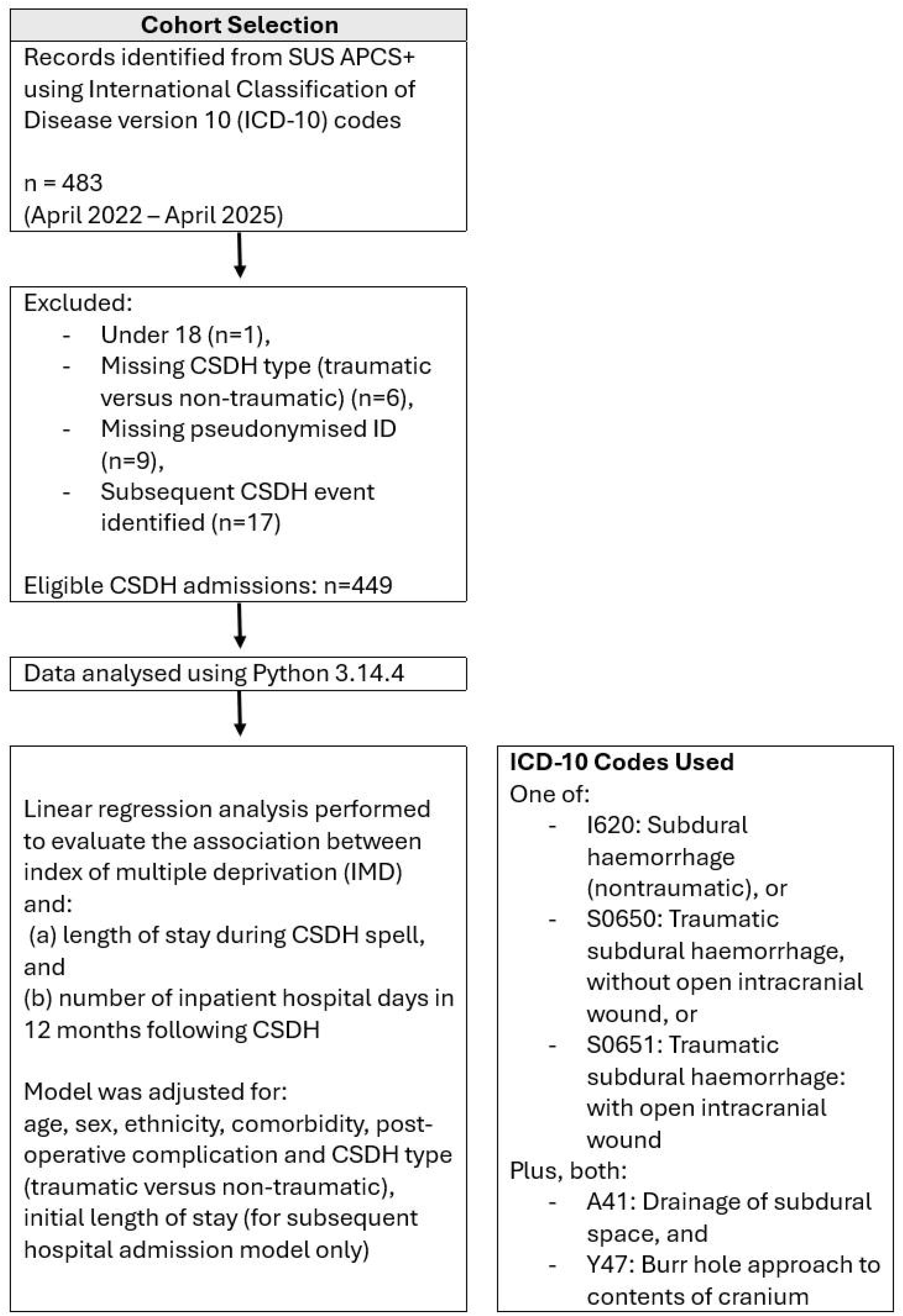
Cohort selection and analysis workflow.

### Outcome and Exposure Measures

The following clinical and sociodemographic metrics were gathered: age, sex, ethnicity (White, Black British/Black Welsh/African/Caribbean, Asian/Asian British/Asian Welsh, mixed/other ethnic groups), index of multiple deprivation (IMD), CSDH aetiology (traumatic or non-traumatic), presence of one or more post-operative complication (hospital acquired pneumonia (HAP), aspiration pneumonia, urinary tract infection (UTI), delirium, surgical wound infection, seizures, stroke, deep vein thrombosis (DVT) or pulmonary embolism (PE)), comorbidity index (see below), length of initial CSDH admission (both overall, and in critical care), death during initial CSDH admission, number of days admitted to hospital in both the preceding and subsequent 12 months, and the admission specialty for this subsequent hospitalisation.

Area-level IMD scores indicate relative levels of neighbourhood socioeconomic deprivation, and are produced by the UK government. They are based on seven independent indices of deprivation: income, employment, education skills and training, health and disability, crime, barriers to housing and services, and living environment deprivation [22]. IMD scores were divided into quintiles, where a score of 1 represents the least socioeconomically deprived and 5 the most socioeconomically deprived.

Comorbidity index was calculated according to the Charlson Comorbidity Index (CCI) [31], but has been modified due to limitations in our ability to accurately identify disease severity for the following comorbidities: liver disease, diabetes mellitus and chronic kidney disease (CKD). For liver disease and diabetes mellitus, we used the lower of two possible scores, assuming less severe disease. For CKD, any diagnosis was given 2 points (where in the CCI, moderate to severe CKD is scored 2 points and mild is scored no points). Patients with one or more of these diagnoses may therefore have had a calculated comorbidity index up to 3 points lower or 2 points higher than their true CCI.

### Data Analyses

Demographic data were presented as absolute counts and percentages for categorical measures, and medians and interquartile ranges (IQRs) for continuous measures. Cohort ethnicity and IMD were compared to 2021 local population census data (the latest available) using standardised mean differences (SMD), which can be found in Supplement Table S1.

Multivariable linear regression analyses were used to test two primary hypotheses; that initial CSDH length of stay (LoS) would be longer for those with greater neighbourhood-level socioeconomic deprivation, and that the number of days spent admitted to hospital in the 12 months after CSDH would be higher for those with greater neighbourhood-level socioeconomic deprivation. Both models include age, sex, comorbidity index, presence of one or more post-operative complications and CSDH aetiology (traumatic versus non-traumatic) as covariates. The second model also included initial hospital LoS. Covariates for both models were selected *a priori* according to data availability and likely impact on the outcome based on current clinical understanding. Unadjusted and adjusted linear regression model outcomes are presented. A p-value<0.05 was considered statistically significant. Power analyses and model assumption tests can be found in Supplement Tables S2 and S3. Both regression models were conducted using heteroscedasticity-consistent (HC3) covariance estimators due to relatively small categorical sub-groups [32]. The untransformed model evaluating the number of days admitted to hospital 12 months after CSDH demonstrated non-linearity; to resolve this assumption violation, a natural log-plus-one transformation was used. The untransformed model is presented in Supplement Figure S1.

Post-hoc analyses were conducted to test the association between IMD, ethnicity and other regression model covariates, detailed in Supplement Table S4.

All analyses were conducted using Python version 3.14.4. No informed patient consent was required, as the dataset was created using routinely collected data during clinical practice. The study does not meet criteria for human subject research according to the NHS Health Research Authority.

## RESULTS

### Baseline Characteristics of Patients with CSDH

449 patients were included in the final analyses (Figure 1). 328 (73.1%) were male, and the median age was 77 years (interquartile range (IQR) 67 years – 85 years) (Table 1). 283 (63.0%) were of white ethnicity, 37 (8.2%) of black ethnicity and 50 (11.1%) of Asian ethnicity. Compared to the local general population, people of black, Asian and minority ethnic backgrounds in this cohort were overrepresented, whereas those of white ethnicity were underrepresented (Supplement Table 1). 346 (77.1%) had at least one comorbidity, most commonly hypertension (53.2% patients) and diabetes (25.2% patients). Most patients were in the least deprived IMD quintile (37.2% versus 5.6% in the most deprived group), these proportions reflect the composition of the local population (Supplement Table S1).

**Table 1:** Baseline cohort characteristics.

|  | **Overall Cohort**<br>N = 449 |
| --- | --- |
| Median age, years (IQR) | 77 (67 – 85) |
| Male sex (%) | 328 (73.1) |
| Ethnicity (%) |  |
| White | 283 (63.0) |
| Asian | 50 (11.1) |
| Black | 37 (8.2) |
| Other | 57 (12.7) |
| Unknown | 22 (4.9) |
| ≥1 comorbidity*, n (%) | 346 (77.1) |
| Index of multiple deprivation quintile, n (%) |  |
| 1 (least deprived) | 167 (37.2) |
| 2 | 109 (24.3) |
| 3 | 63 (14.0) |
| 4 | 57 (12.7) |
| 5 (most deprived) | 25 (5.6) |
| Unknown | 28 (6.2) |
| Median CSDH spell length of stay, days (IQR) | 7 (4 – 12) |
| Traumatic CSDH aetiology (%) | 237 (52.8) |
| ≥1 post-operative complication<sup>†</sup> during CSDH spell, n (%) | 120 (26.7) |
| Died during CSDH spell, n (%) | 5 (1.1) |
| Median time spent in hospital in the 12 months before CSDH, days (IQR) | 0 (0 – 8) |
| Median time spent in hospital in the 12 months after CSDH, days (IQR) | 5 (0 – 28) |

\*Diabetes, hypertension, dementia, chronic kidney disease (CKD), liver disease, previous stroke, previous transient ischemic attack, previous myocardial infarction, peripheral vascular disease, chronic pulmonary disease, connective tissue disease, peptic ulcer disease, hemiplegia, malignancy, or acquired immunodeficiency syndrome.
<sup>†</sup>Delirium, urinary tract infection, hospital acquired pneumonia, aspiration pneumonia, surgical wound infection, pulmonary embolism, deep vein thrombosis, seizures, or stroke.

Around half (52.8%) of patients had traumatic CSDH aetiology (Table 1). Median length of stay during the initial CSDH spell was 7 days (IQR 4 – 12). 5 (1.1%) patients died during initial admission, and 120 (26.7%) had at least one post-operative complication. In the 12 months after initial CSDH, the median time admitted to hospital for any reason was 5 days (IQR 0 – 28), versus 0 days (IQR 0 – 8) in the 12 months prior.

### Association Between Covariates and CSDH Admission Length of Stay

No significant association was found between initial spell LoS and IMD quintile (Kruskal-Wallis H-statistic = 7.52, p=0.111) or ethnicity (Kruskal-Wallis H-statistic = 9.42, p=0.051). We note that relatively small ethnicity subgroup sizes limit the power to detect a significant difference.

A robust multivariable linear regression model was used to assess the impact of clinical and sociodemographic factors on the initial spell LoS. Higher comorbidity index (β=1.24, 95%CI 0.28 to 2.20), having at least one post-operative complication (β=8.89, 95%CI 4.00 to 13.78), male sex (β=3.90, 95%CI 1.11 to 6.69), and traumatic aetiology (β=4.51, 95%CI 1.13 to 7.89) were independently associated with increased LoS (Figure 2). IMD and ethnicity were not independently associated with the initial CSDH length of stay. The model power was 0.79.

**Figure 2:**
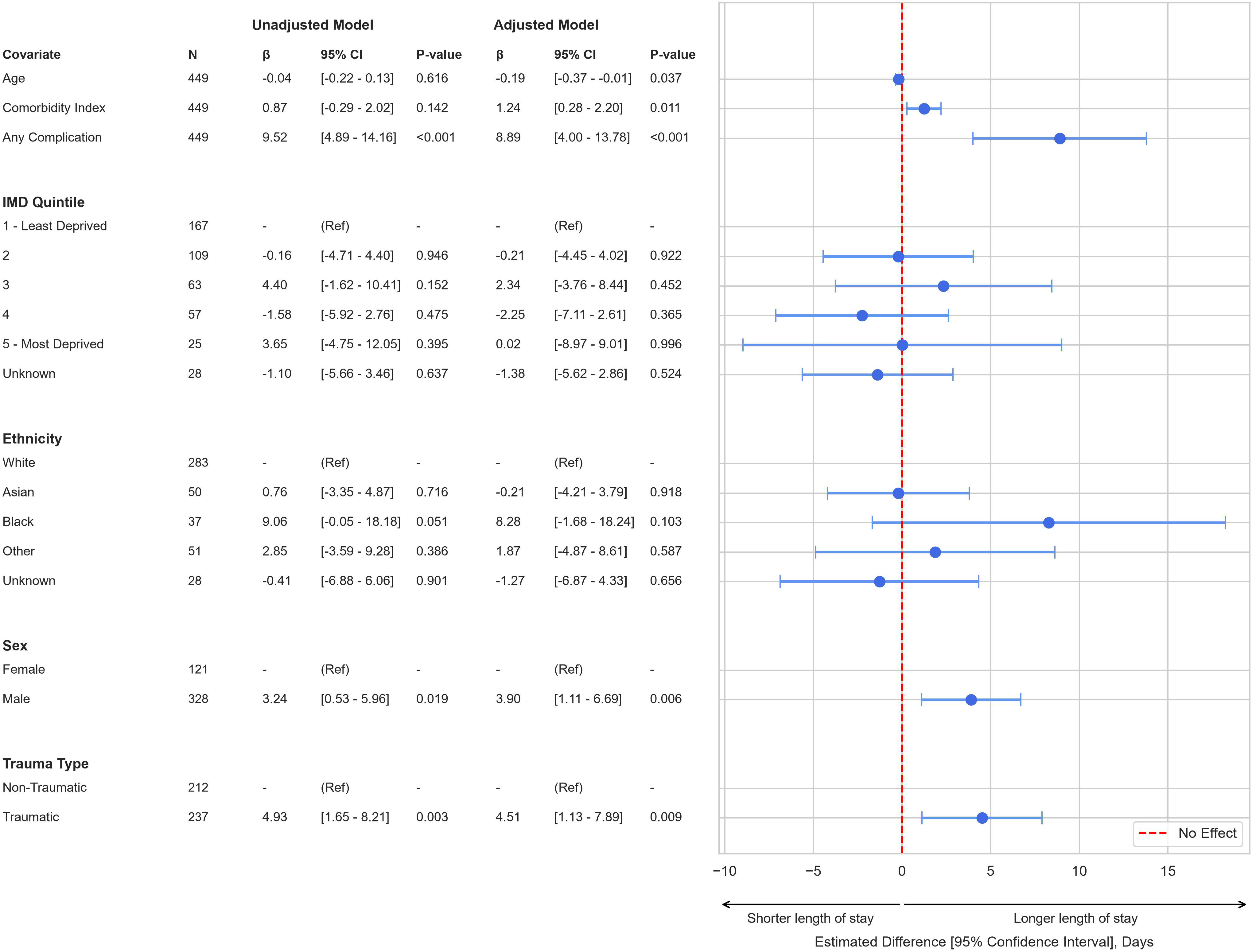
Multivariable linear regression, using heteroscedasticity-consistent (HC3) covariance estimator, to show the effect of clinical and sociodemographic variables on hospital length of stay for chronic subdural haematoma (CSDH).

### Association Between Covariates and Inpatient Hospital Days in the 12 Months After CSDH Admission

For the most deprived patients in IMD quintile 5, the median time spent in hospital 12 months before CSDH was 8 days (IQR 0 - 28), compared to 0 days (IQR 0 - 5.5) for the least deprived in IMD quintile 1 (Figure 3). During the 12 months after CSDH, median time spent in hospital by the most deprived patients was 26 days (IQR 1 - 48) versus 4 days (IQR 0 - 18.5) for the least deprived. These differences are statistically significant (Kruskal-Wallis H-statistic = 16.8, p=0.004; Dunn’s post-hoc p-value = 0.043 between the most and least deprived quintiles).

**Figure 3:**
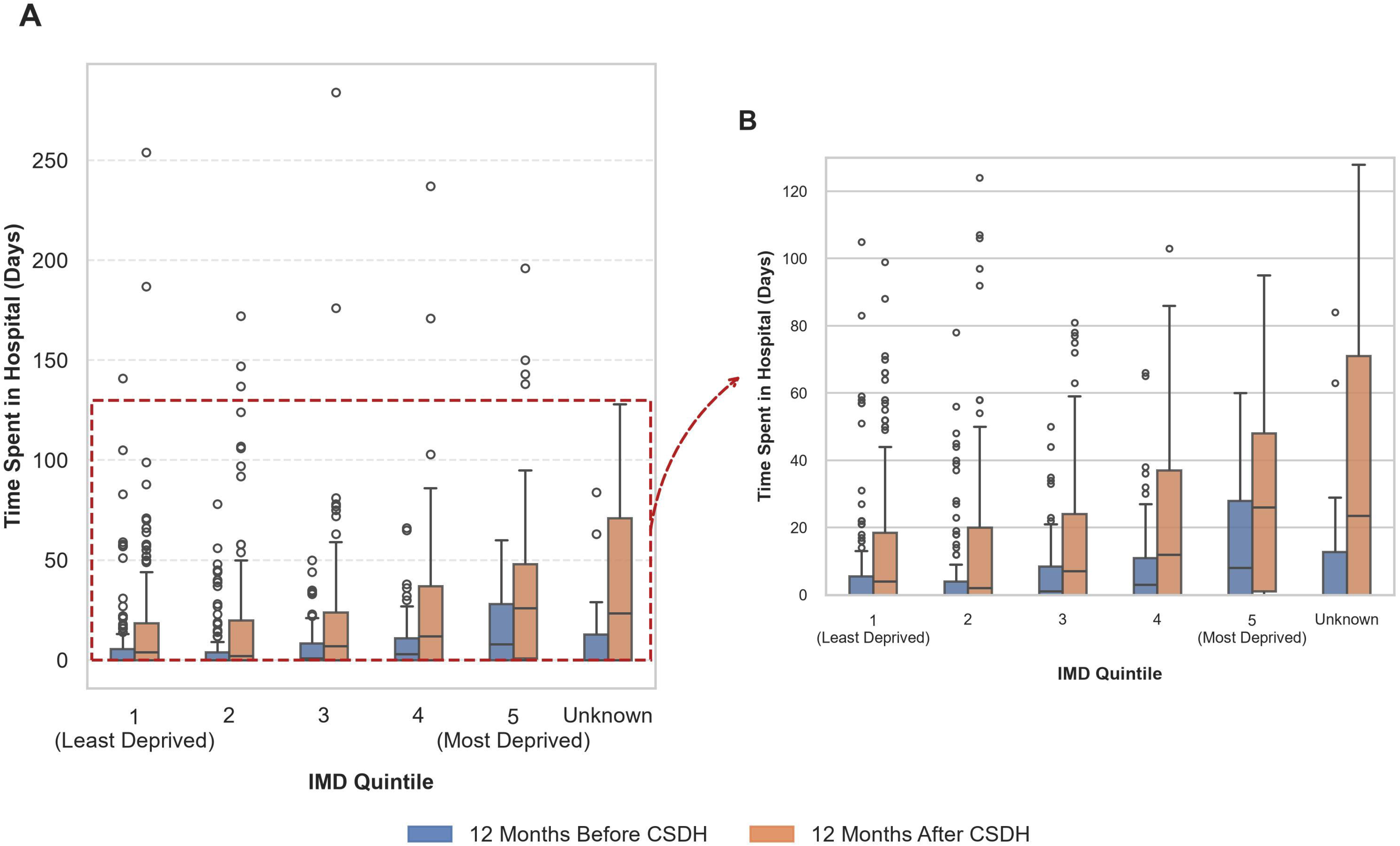
Box plot of median (IQR) inpatient days in hospital 12 months before and 12 months after chronic subdural hematoma (CSDH) by index of multiple deprivation (IMD). (A) Complete data. (B) Y-axis truncated at 130 days.

The median change before and after CSDH was 17 days (IQR 0 - 28) for the most deprived, and 0 days (IQR -2 - 14) for the least deprived. Spearman rank correlation showed a negative correlation in the median change in number of days spent in hospital by IMD quintile (*p* = -0.11, p=0.019).

A natural log-transformed multivariable linear regression model (power = 1.0) showed a graded association between socioeconomic deprivation and time spent in hospital in the year after CSDH. Those in the most deprived groups spent the longest in hospital. This association was robust to model adjustments for the other clinical and demographic factors included (Figure 4). The most deprived patients were estimated to spend almost 4.5-fold longer per year in hospital compared to the least deprived (fold change = 4.45, 95%CI 2.20 to 9.01). In the untransformed model (Supplement Figure S1), the estimated number of days spent in hospital by the most deprived group was 31 days more than the least deprived group (β=31.86, 95%CI 15.95-47.77).

**Figure 4:**
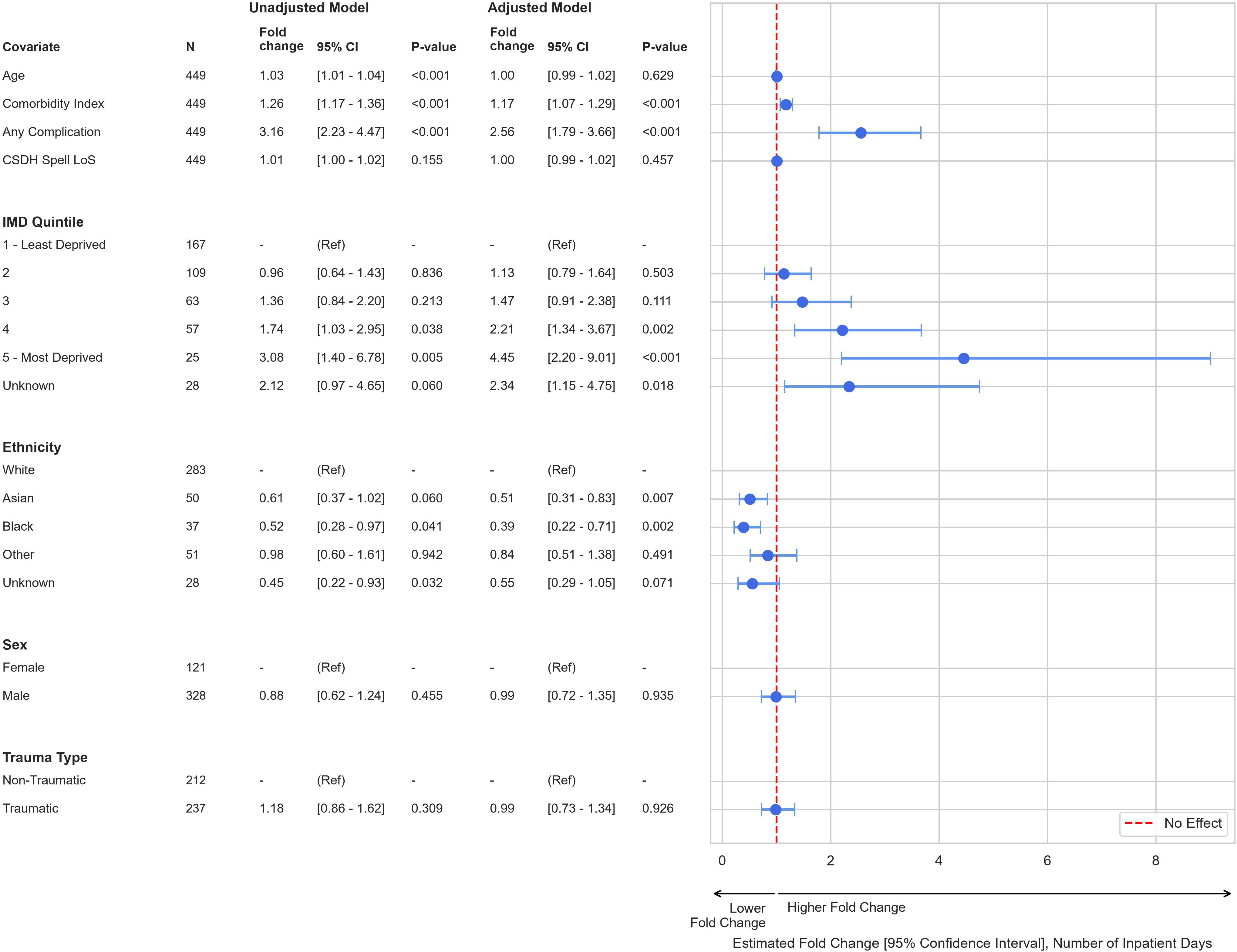
Natural log-transformed multivariable linear regression, using heteroscedasticity-consistent (HC3) covariance estimator, to show the effect of clinical and sociodemographic variables on the fold-change in number of inpatient days during the 12 months following chronic subdural haematoma (CSDH).

In the adjusted model the following factors were also associated with more inpatient hospital days the year after CSDH: higher comorbidity index (fold change = 1.17, 95%CI 1.07 to 1.29) and having at least one post-operative complication (fold change = 2.56, 95%CI 1.79 to 3.66) (Figure 4). Asian and Black ethnicity were associated with less time spent in hospital during the 12 months after CSDH (fold change = 0.51, 95%CI 0.31 to 0.83, and fold change = 0.39, 95%CI 0.22 to 0.71 respectively).

Of the admissions during the 12 months after CSDH, 917 (82.8%) were to a medical specialty. For all IMD quintiles, medical specialties were the most common admission type (Figure 5).

**Figure 5:**
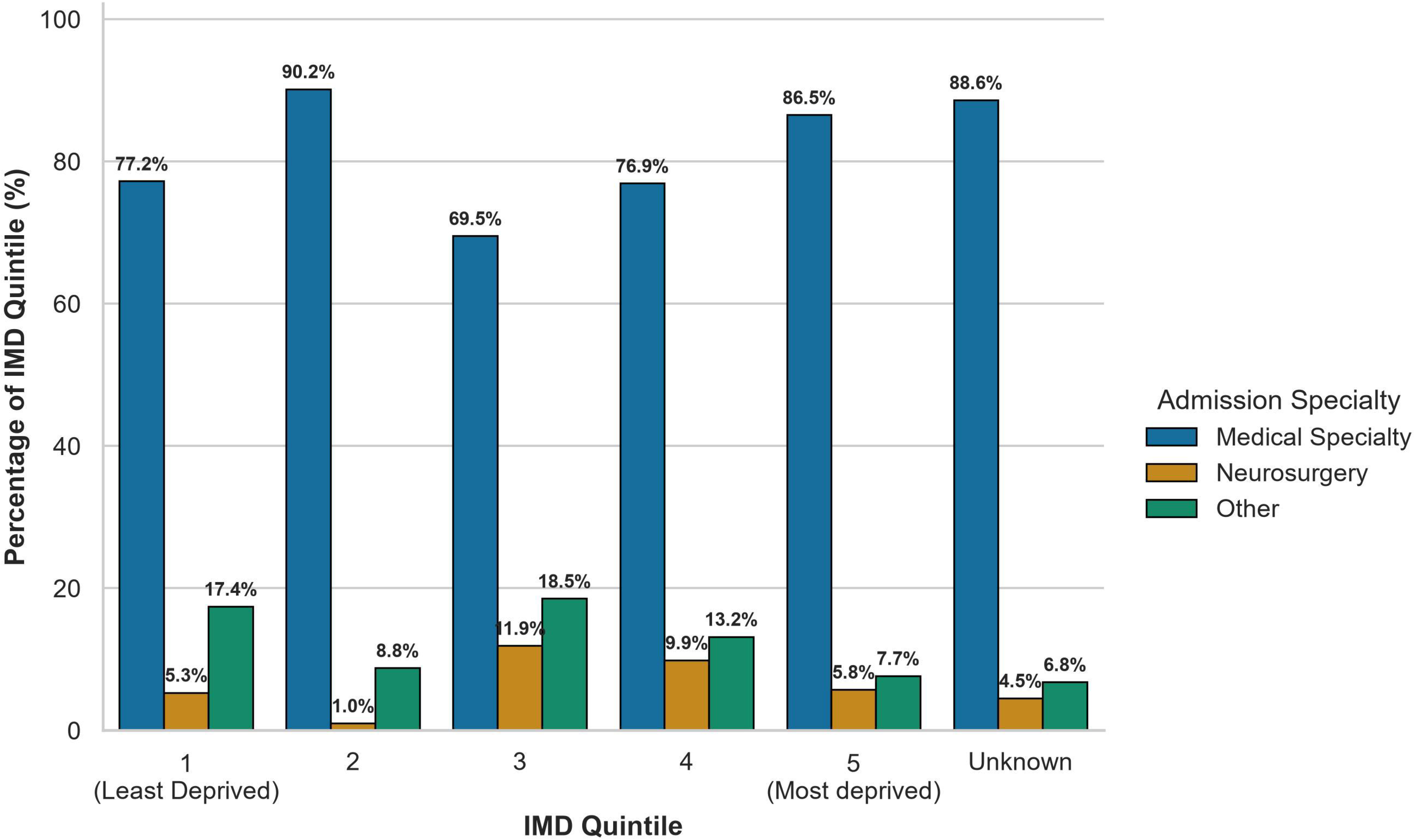
Admission specialties for any admission during the 12 months after chronic subdural haematoma (CSDH) by IMD Quintile.

### Post-Hoc Analyses

Post-hoc analyses were conducted to assess the relationship between IMD, ethnicity and other covariates (Supplement Table S4). Neighbourhood-level deprivation varied by ethnicity (Kruskal-Wallis H-value = 28.67, p<0.001). Dunn’s post-hoc analyses identified that those of black and Asian ethnicity experienced greater neighbourhood-level deprivation than those of white ethnicity (median IMD 4 (IQR 3 to 5) versus IMD 3 (IQR 2 to 4), p<0.001 and median IMD 4 (IQR 3 to 5) versus IMD 4 (IQR 2 to 4), p=0.028, respectively). Similarly, age varied by ethnicity (Kruskal-Wallis H=21.89, p<0.001). Black patients were significantly younger than white patients (median age 69 years versus 79 years, Dunn’s test p=0.008). There was, however, no association between ethnicity and having one or more post-operative complication, co-morbidity index, traumatic versus non-traumatic aetiology or length of critical care stay. There was also no association between IMD quintile and age, having one or more post-operative complication, co-morbidity index, traumatic versus non-traumatic aetiology or critical care length of stay.

## DISCUSSION

To our knowledge, this is the first UK study investigating the association between socioeconomic deprivation and outcomes after CSDH. We show that in a diverse cohort of patients, neighbourhood-level deprivation has no significant impact on LoS during the acute admission, but does significantly increase time spent hospitalised during the subsequent 12 months.

Even within a universal healthcare system such as the NHS, socioeconomic deprivation is associated with poorer health outcomes, driven by a complex network of factors including individual behaviours, environmental exposures, access to care, comorbid disease and wider social determinants of health [21]. In our cohort, hospital usage was already higher amongst those in the most deprived groups, which is unsurprising and consistent with findings reported in the current literature [21,33,34]. We show, however, that CSDH widens this gap in healthcare usage. While IMD had no significant independent effect on initial LoS, those in the most deprived quintile were estimated to spend around 4.5 times longer in hospital the year after CSDH than the least deprived group (fold change = 4.45, 95%CI 2.20 to 9.01), which, in the untransformed model, equated to around a month longer in hospital. This was after adjusting for age, ethnicity, CSDH aetiology, comorbidities, post-operative complications and initial hospital length of stay. Our data support previously reported findings that CSDH is a critical sentinel health event [13,35], and, additionally indicate that the negative impacts are inequitably experienced more by those who are the most socioeconomically deprived. This indicates a socioeconomic health inequality in this cohort, defined as ‘avoidable, unfair and systematic differences in health between different groups of people’ [36]. As age and socioeconomic disadvantage are the two strongest risk factors for avoidable hospital admission [37], we believe that these are important findings not only from a health inequalities perspective, but also with respect to rising healthcare costs.

Future work in this area should focus on understanding the factors mediating the association between deprivation and outcomes after CSDH. This may include large, prospective multicentre studies, which could additionally investigate the effect of individual-level deprivation. We suggest that studies designed to elucidate the underlying mechanisms of these findings would also be worthwhile; in particular, understanding the effect of baseline frailty, alcohol and substance misuse, pre-admission medications and CSDH characteristics would be enlightening. It would also be of considerable interest to understand the impact of deprivation on outcomes in conservatively managed CSDH patients.

More than 80% of patients admitted to hospital during the 12 months after CSDH were admitted under medical specialities. Future work to understand the main causes of these admissions would be worthwhile, and could help to shape community services aiming to prevent readmission after CSDH. This would reduce the burden on medical wards by these patients. Any services developed must be accessible for all, to minimise ongoing health inequities.

Our results also indicate possible inequalities in outcomes between white and non-white ethnicity; notably, that black and Asian patients are estimated to spend less time in hospital during the year after CSDH, despite the fact that greater neighbourhood-level deprivation was experienced by this group. The relationship between ethnicity and socioeconomic status is complex [38], and these findings may be affected by factors we were unable to capture, such as barriers in access to care, CSDH characteristics or differences in prescribed medications. There is also the potential that differences in cultural-social factors, such as family proximity, multi-generational living or care giving expectations are protective against hospital admissions. We suggest that these findings warrant further study to maximise equity of care.

We report these findings at a time of considerable multi-disciplinary interest for CSDH research in the UK. Stubbs and colleagues recently published a seminal expert consensus guideline, outlining recommendations for best practice in the care of patients with CSDH [39]. In line with this, efforts are underway within our local neurosurgical network to introduce a package of interventions aiming to improve, standardise and ensure equitable care for CSDH patients in South West London. We acknowledge the substantial work being performed by this group and the Improving Care in Elderly Neurosurgery Initiative (ICENI) working group, and hope that these results will add to the current body of work driving improvements in, and standardisation of, care for this large and often vulnerable patient group.

## Limitations

There are several limitations to this study. Results are based on administrative data which depend on accurate and consistent clinical coding. IMD is an England-specific measure, and our cohort is derived from a single centre, which limits the overall generalisability of our results. Because IMD is an area-based measure of relative deprivation there is also likely to be substantial variation in individual-level deprivation within each IMD quintile. Due to limited data availability we were unable to adjust for potentially important confounders or mediators, including hematoma characteristics, medications, alcohol or substance use, and frailty or other functional baseline status. Collecting these in future prospective studies may reveal the underlying mechanisms driving our observed differences, and would help to target interventions aiming to reduce readmission after CSDH amongst the most socioeconomically deprived. We were unable to fully calculate the Charlson Comorbidity Index due to a limited ability to determine comorbid disease severity. Due to relatively small sub-group sample sizes, linear regression estimates should be interpreted with caution. Larger, multi-centre studies are warranted to replicate our findings. Finally, as these analyses are observational, no causal effect can be interpreted, and the findings should be understood to describe inequalities in CSDH outcomes within this cohort from Southwest London.

## CONCLUSION

This is the first UK study which demonstrates socioeconomic health inequalities in CSDH outcomes; higher neighbourhood-levels of deprivation are associated with longer time spent hospitalised in the year following CSDH. There is a need both to gain greater insight into the causal or mediating factors in this association, and for work to improve equity of outcomes after CSDH.

## Data Availability

All data produced in the present study are available upon reasonable request to the authors.

## ETHICS STATEMENTS

Institutional board approval from St George’s University Hospitals NHS Foundation Trust was provided as a review of services (AUDI004846 on 1/5/2026).

### Patient consent for publication

Not applicable.

## CONTRIBUTORS

**EC:** conceptualisation (equal); methodology (equal); formal analysis (lead); visualisation (lead); writing – original draft (lead); writing – review and editing (equal). **MF:** writing – review and editing (equal), supervision. **CR:** data curation (lead). **KB:** writing – review and editing (equal). **JBT:** writing – review and editing (equal). **MH**: conceptualisation (equal); methodology (equal); writing – review and editing (equal); supervision (lead). EC is the guarantor and accepts responsibility for the overall content of the work.

## FUNDING

**KB** receives funding from the Epilepsy Research Institute UK (Grant: F2201 Bush). No other funding to report in relation to the presented manuscript.

## COMPETING INTERESTS

None to declare.

## Notes

### Competing Interest Statement

The authors have declared no competing interest.

