## Supplement for "Beyond the Haematoma: Socioeconomic Deprivation and Outcomes in Chronic Subdural Hematoma in a Single-Centre Cohort Study"

**Supplementary File**

**Baseline Characteristics**

**Table S1:** Comparison of cohort ethnicity and Index of Multiple Deprivation to 2021 local census data

|  | **Cohort Count (%)** | **Census Proportion** | **Standardised Mean Difference (SMD)** | **Chi^2^ Test (p-value)** |
| --- | --- | --- | --- | --- |
| **IMD Quintile** |  |  |  |  |
| 1 (Least deprived) | 167 (39.7%) | 37.6% | 0.0428 | Χ^2^ = 7.97 (p=0.093) |
| 2 | 109 (25.9%) | 24.5% | 0.0312 |  |
| 3 | 63 (15.0%) | 20.0% | -0.1335 |  |
| 4 | 57 (13.5%) | 13.3% | 0.0070 |  |
| 5 (Most deprived) | 25 (5.9%) | 4.5% | 0.0620 |  |
| **Ethnicity** |  |  |  |  |
| White | 240 (65.2%) | 81.9% | -0.3842 | Χ^2^ = 233.41 (p<0.001) |
| Black | 31 (8.4%) | 4.3% | 0.1677 |  |
| Asian | 46 (12.5%) | 8.3% | 0.1386 |  |
| Mixed or Multiple Ethnic Group | 6 (1.6%) | 3.5% | -0.1205 |  |
| Other | 45 (12.2%) | 2.0% | 0.4071 |  |

**Multivariate Linear Regression Model Assumption Tests**

**Table S2:** Assumption test outcomes for the multivariable linear regression model analysing initial spell length of stay.

| **Assumption** | **Test** | **Outcome** | **Solution** |
| --- | --- | --- | --- |
| Linearity | Visual inspection (residuals vs. fitted plot) | Pass | - |
| Independence | Durbin-Watson | Durbin-Watson Statistic = 2.25 (pass) | - |
| Heteroscedasticity | Bresusch-Pagan | P-value = 0.136 (pass) | Heteroscedasticity-consistent (HC3) method used across both models. |
| Normality of Residuals | Shapiro-Wilk | P<0.0001 (violated) | N>100; safe to proceed with no model amendments. |
| Multicollinearity | Variance Inflation Factor (VIF) | All 1<VIF<5 | - |

**Table S3:** Assumption test outcomes for the multivariate linear regression model analysing number of days spent in hospital in the 12 months after CSDH.

| **Assumption** | **Test** | **Outcome** | **Solution** |
| --- | --- | --- | --- |
| Linearity | Visual inspection (residuals vs. fitted plot) | Fail | Natural log-plus-one transformation |
| Independence | Durbin-Watson | Durbin-Watson Statistic = 2.05 (pass) | - |
| Heteroskedasticity | Bresusch-Pagan | P-value = 0.079 (pass) | Heteroscedasticity-consistent (HC3) method used across both models. |
| Normality of Residuals | Shapiro-Wilk | P<0.0001 (violated) | N>100; safe to proceed with no model amendments. |
| Multicollinearity | Variance Inflation Factor (VIF) | All 1<VIF<5 | - |

**Untransformed Linear Regression Model – Number of Days Spent in Hospital in the 12 Months After CSDH**

**Figure S1:** Untransformed multivariable linear regression to show the effect of clinical and sociodemographic variables on the number of days spent hospitalised in the 12 months after chronic subdural hematoma.

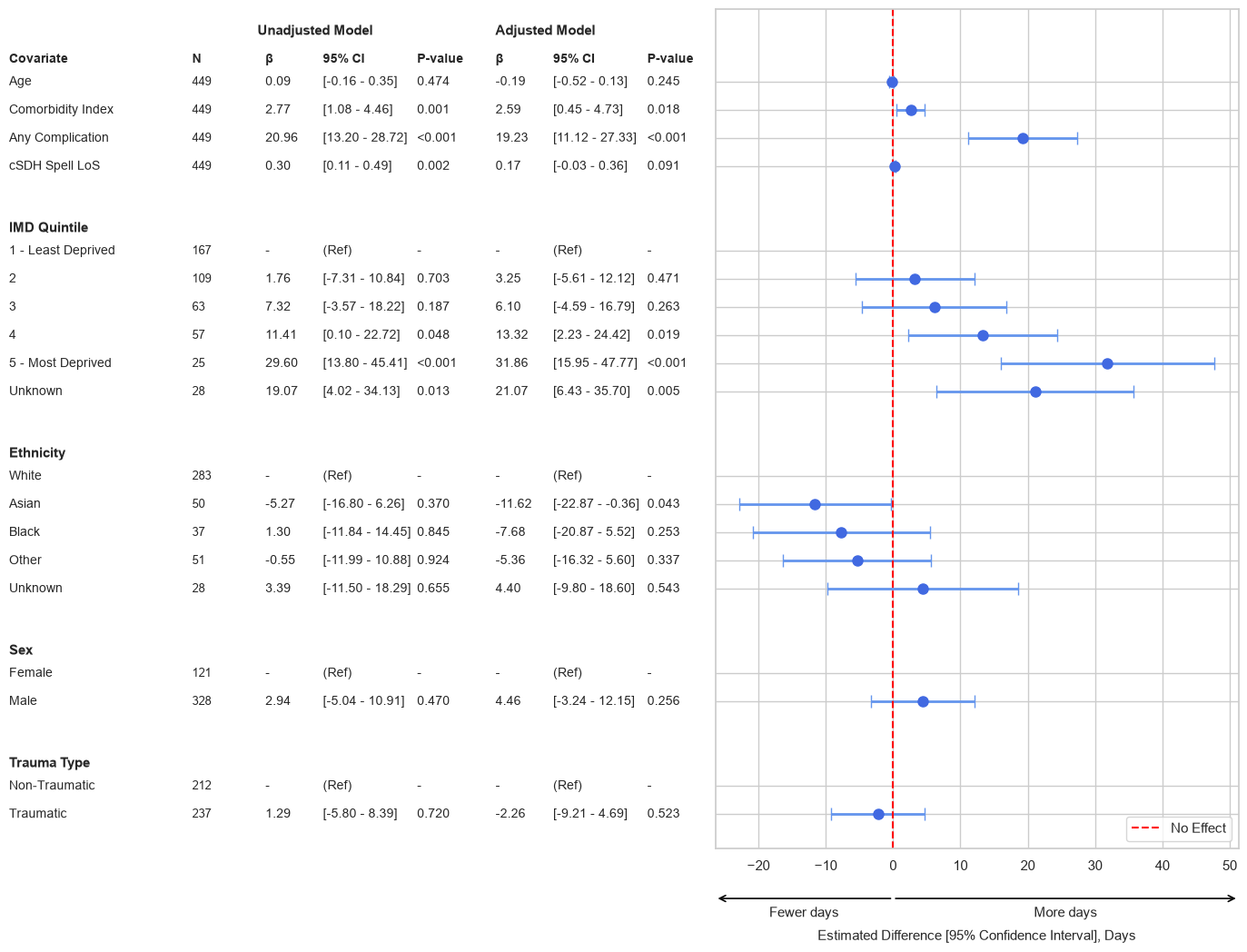

**Post-Hoc Analyses**

**Table S4:** Post-hoc analyses testing the association between ethnicity, Index of Multiple Deprivation and other covariates

|  | Test | Effect Size | P-value |
| --- | --- | --- | --- |
| Association with ethnicity | | | |
| Index of Multiple Deprivation | **Kruskal-Wallis** | **H=21.89** | **<0.001** |
| Age | **Kruskal-Wallis** | **H=19.42** | **<0.001** |
| Presence of ≥1 post-operative complication | Chi-Square | Χ^2^=2.07 | 0.72 |
| Comorbidity Index | One-Way ANOVA | F=1.68 | 0.15 |
| Traumatic versus non-traumatic CSDH aetiology | Chi-Square | Χ^2^=6.07 | 0.19 |
| Length of critical care stay | Kruskal-Wallis | H=5.11 | 0.28 |
| Association with Index of Multiple Deprivation | | | |
| Age | Kruskal-Wallis | H=9.71 | 0.05 |
| Presence of ≥1 post-operative complication | Chi-Square | Χ^2^=1.11 | 0.89 |
| Comorbidity Index | Kruskal-Wallis | H=4.74 | 0.31 |
| Traumatic versus non-traumatic CSDH aetiology | Chi-Square | Χ^2^=4.28 | 0.37 |
| Length of critical care stay | Kruskal-Wallis | H=3.40 | 0.49 |
